# Circadian biomarker signatures for differentiating unipolar from bipolar depression

**DOI:** 10.1101/2025.04.07.25325364

**Authors:** Xin-Ling Wang, Lei Huang, Jiashu Yao, Yanhua Qin, Keming Ren, Yuedi Shen, Wei Chen

**Author notes:** Corresponding authors: Wei Chen, (W. Chen) and Yuedi Shen, (Y. Shen).

## Abstract

**Importance:** Major depressive disorder (MDD) and bipolar disorder (BD) are often misdiagnosed during depressive episodes, therefore, exploring biomarkers for differential diagnosis is important.

**Objective:** To identify circadian biomarker signatures in patients’ peripheral blood that differentiate MDD from BD during depressive states.

**Design, setting, and participants:** This case-control study recruited patients with MDD and BD at depressive state diagnosed by the Diagnostic and Statistical Manual of Mental Disorders Fifth Edition (DSM-5) and Health control (HC) subjects from January 2021 to May 2023. We collected serum samples to detect levels of Period (PER)1/PER2/phosphorylated cAMP reaction element binding protein (pCREB).

**Main outcomes and measures:** Participants’ clinical data were evaluated by HAMD-17 and MDQ scales. Blood samples (*n* = 100) were collected and extracted for serum followed by detecting serum PER1, PER2 and pCREB levels by ELISA.

**Results:** There were 100 participants in the cohort, including 40 in the MDD group, 30 in the BD group and 30 in the health control (HC) group. 71% were female in the cohort. The mean (SD) serum PER1 level in the HC group was 12.05 (2.96) ng/mL, in the MDD group was 8.41 (2.96) ng/mL, significantly decreased vs the HCs, and in the BPD group that was 16.05 (3.60) ng/mL, significantly increased vs the HCs (adjusted *P* < 0.0001). The serum pCREB level in the HC group was 205.2 (49.57) ng/mL, in the MDD group was 192.6 (38.52) ng/mL, and that in the BPD group was 290.0 (72.10) ng/mL, which was significantly increased vs the HCs and vs the MDD group. For the differential diagnosis between MDD and BPD, PER1 & pCREB (AUC, 0.9858) show similar high diagnostic efficiency as combined biomarkers of PER1, PER2 & pCREB (AUC, 0.9906).

**Conclusions and relevance:** This study reveals the importance of serum PER1, combined use of serum PER1 and pCREB as well as that of serum pCREB, PER1 and PER2 in differentiating MDD from BPD.

**Key Points:** *Question:* Can major depressive disorder (MDD) be distinguished from bipolar disorder (BD) during their depressive episodes by circadian biomarkers in patient’s peripheral blood?

*Findings:* In this case-control study of 100 participants, serum PER1 levels were significantly decreased in the MDD group but were significantly increased in the BPD group, serum PER2 levels were significantly elevated in both MDD and BPD groups, and serum pCREB levels were significantly increased in the BPD group, compared with the HC group. Besides, In the MDD group, serum PER1 levels were significantly positively associated with mood disorder questionnaire (MDQ) scores; and serum pCREB levels were significantly negatively associated with MDQ scores. In the BPD group, serum pCREB levels were significantly negatively associated with HAMD-17 scores. The combined use of serum pCREB and PER1 and the three biomarkers (pCREB, PER1 and PER2) had similar diagnostic values in differentiating MDD from BPD.

*Meaning:* The serum PER1 has great value for differentiating MDD from BPD, and combined use of serum biomarkers of pCREB/PER1 or pCREB/PER1/PER2 shows higher efficacy in differentiating MDD from BPD.

## Introduction

Major depressive disorder (MDD) and bipolar disorder (BD) are common mood disorders. MDD is a disease with recurrent unipolar depressive episodes (UPD). In contrast, BD is a disease characterized by recurrent manic and depressive episodes. As they exhibit overlapping systems during the depressive episodes, patients with MDD and BD are often misdiagnosed ^1–5^. At present, the differential diagnosis between MDD and BD mainly relies on clinical symptoms ^6^, and the misdiagnosis rate of them is high in the early stage ^7^. Therefore, identifying sensitive and specific biomarkers may be valuable for differentiating MDD from bipolar depression (BPD) ^8^.

A large body of evidence has implicated circadian rhythm in the pathophysiology and treatment of mood disorders ^9–19^. Among the core circadian genes, *Per1* and *Per2* have been implicated involved in mood disorders ^16, 20–27^, and *Per1* and *Per2* have an identical sequence (TGACGTCA) that is the cAMP response element (CRE) site, the binding site of phosphorylated cAMP reaction element binding protein (pCREB), while *Per3* and other core circadian genes don’t have this site. Further, pCREB a transcription factor which regulates the genes containing CRE site ^28^. Moreover, pCREB is the convergent downstream molecule of several cellular pathways involved in mood regulation, including mitogen-activated protein kinases (MAPKs), calcium, protein kinase A (PKA), and protein kinase C (PKC) ^29, 30^, etc. Furthermore, pCREB and CREB have also been widely reported to be involved in mood disorders ^30–36^. However, so far, it has not been demonstrated whether circadian biomarkers can be used in the differential diagnosis between MDD and BD. Therefore, in this study, we focused on pCREB, PER1 and PER2.

Here, we conducted a case-control study using serum samples from (*n* = 100) subjects with MDD (*n* = 40) and BPD (*n* = 30) compared with HCs (*n* = 30). By one-way ANOVA, we compared serum levels of PER1, PER2 and pCREB levels among the MDD, BPD and HC groups. Next, the correlation between clinical symptoms assessed by MDQ and HAMD-17 and these serum biomarkers were analyzed. Finally, we explored the model to distinguish MDD from BPD using separate or combined serum biomarkers.

## Methods

This study was approved by the Ethics Committee of Sir Run Run Shaw Hospital, Zhejiang University School of Medicine (No. 20191203-13). All study participants were informed of the purpose and methods of the study, and each participant provided informed consent before enrolment.

### Study Participants

The study enrolled patients hospitalized for MDD and BPD between January 2021 to May 2023, aiming to identify circadian biomarkers among pCREB, PER1 and PER2 able to differentiate MDD from BPD, as well as from HCs.

The inclusion criteria: patients who met the DSM-5 diagnostic criteria for BD or MDD; patients with BD were currently experiencing a depressive episode; 18-65 years old; HAMD-17 scores ≥ 18 points; MDQ scores ≥ 7 in the BD group and < 7 in the MDD group; patients signed the informed consent form. The exclusion criteria: patients with BD who were experiencing a manic, hypomanic, or mixed state; other psychiatric disorders, such as schizophrenia and schizoaffective disorder; pregnant or lactating women; intellectual disability, substance abuse disorder, somatoform disorders, or mood disorder secondary to other diseases.

Inclusion criteria for the control group: age 18-65 years; HAMD-17 < 7; MDQ < 7; signed the informed consent form. Exclusion criteria for the control group: pregnant or lactating women; reported a history of a psychotic disorder, mood disorder, anxiety disorder, substance abuse disorder, brain trauma, or intellectual disability; reported a history of mental illness or serious mental illness in their first-degree relative.

### HAMD scale assessment

The HAMD-17 was used in this study. The scores were categorized as follows: a total score ≤ 7 points indicates no depressive symptoms; a total score ≥ 18 indicates moderate depression; and a total score ≥ 24 indicates more severe depression.

### MDQ scale assessment

The MDQ is the most commonly used scale to screen for BD. Hirschfeld et al. showed that the MDQ with a defined value of 7 had a sensitivity of 0.73 and a specificity of 0.9 for differentiating MDD from BPD ^37^.

### Blood sample collection and ELISA

All participants were fasting at the time of collection, and we collected 5 ml of blood at the same time point (8:00, zeitgeber time (ZT) 2). These samples were collected from the elbow vein and placed in a test tube with anticoagulant. The samples were then centrifuged at 3000 rpm/min for 15 min on the same morning. After centrifugation, the sample was divided into three layers: the upper layer, the intermediate layer and the lower layer. We extracted the upper serum layer, placed it into 2-ml EP tubes, and stored at −80 □. Then we used the human pCREB ELISA kit (Cat No. JM-4980H1), human PER1 ELISA kit (Cat No. JM-1128H1), and human PER2 ELISA kit (Cat No. JM1129H1) to detect the serum levels of pCREB, PER1 and PER2 in each group via ELISA. The protocols were followed to the instructions of the ELISA kits.

### Statistical Analysis

The data were analysed by SPSS 26 software and GraphPad Prism 9.0 software. Age, HAMD-17 scores and MDQ scores were analysed by Welch’s ANOVA with Dunnett’s T3 multiple comparisons test. Education duration was analysed by one-way ANOVA with Tukey’s multiple comparisons test. In addition, serum PER1, PER2 and pCREB levels were analysed by one-way ANOVA with Tukey’s multiple comparisons test and Welch’s ANOVA with Dunnett’s T3 multiple comparisons test according to the normality and homogeneity of variance tests of the data. The correlations of HAMD-17/MDQ scores and serum PER1/PER2/pCREB levels were analyzed by Pearson’s correlation analysis. To assess the respective and combined use of serum PER1/PER2/pCREB in differentiating MDD from BPD, their sensitivity, specificity, Youden index, and area under the ROC curve (AUC) were analyzed via ROC analysis (for respective biomarker), binary (for two combined biomarkers) and multiple regression analysis (for three combined biomarkers). *P* < 0.05 was considered significant.

## Results

### Study Overview

Of 100 participants, 71 (71%) were female, 40 were patients with MDD, 30 were patients with BPD, and 30 were healthy controls. The mean (SD) age was 29.25 (12.91) years in the MDD group, 21.63 (8.95) years in the BD group, and 30.43 (8.52) in the HC group (eTable 1 in Supplement 1). All of the patients were unrelated Chinese. The patients were recruited from the population of inpatients in the Psychiatry department at Sir Run Run Shaw Hospital, Zhejiang University School of Medicine, from January 2021 to May 2023. The research flow chart is shown in Figure 1.

**Figure 1.**
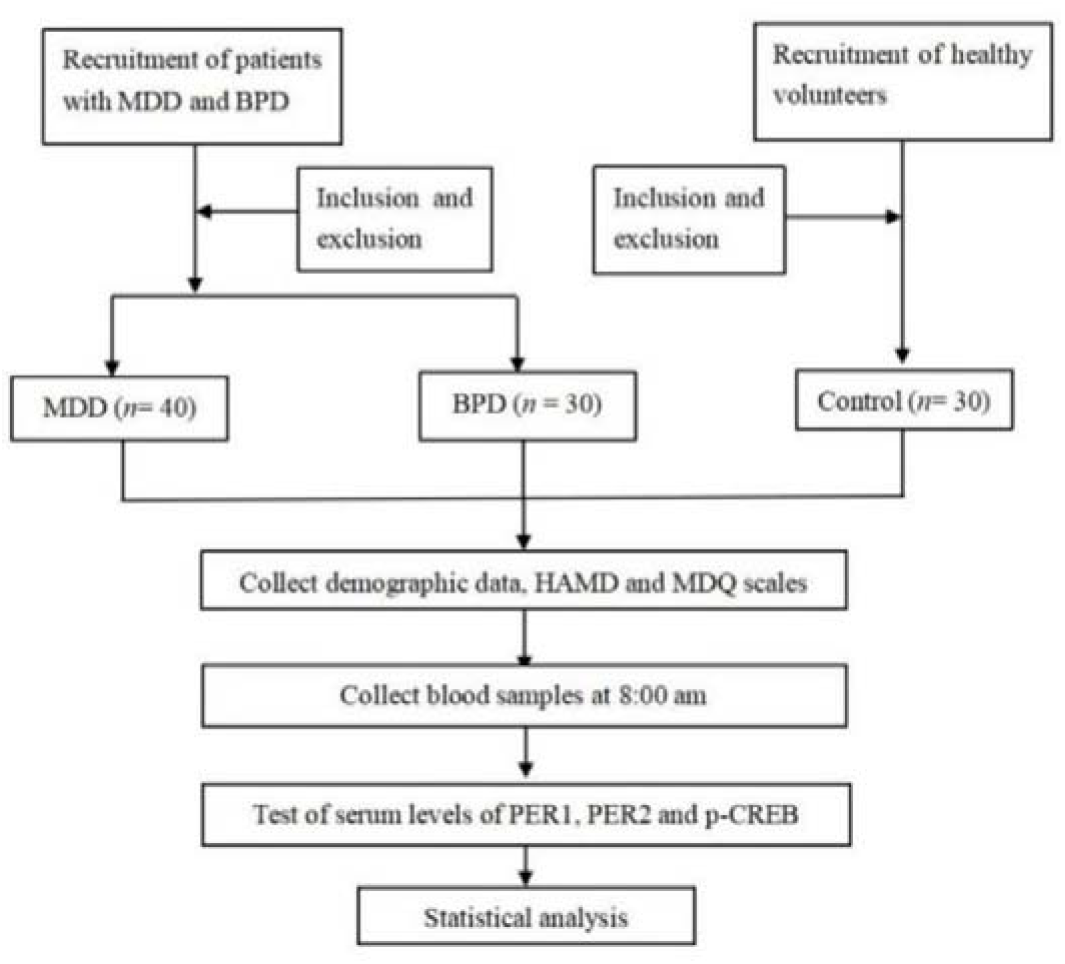
Research flow chart. MDD, major depressive disorder; BPD, bipolar depression; HAMD, Hamilton Depression Scale; MDQ, mood disorder questionnaire.

There were no significant differences in education duration in the MDD and BPD groups, compared with the HC group (*P* = 0.515 and *P* = 0.151 respectively, eTable 1 in Supplement 1). There was also no significant difference in age in the MDD group vs. the HC group (*P* = 0.955, eTable 1 in Supplement 1). However, the age in the BPD group was significantly low vs the HC group (*P* = 0.001) and vs the MDD group (*P* = 0.014, eTable 1 in Supplement 1).

### Comparisons of serum levels of pCREB, PER1 and PER2 in the MDD and BPD groups

We compared the serum levels of pCREB, PER1 and PER2 among all the three groups by one-way ANOVA or Welch’s ANOVA as appropriate (PER1: One-way ANOVA, *F* _(2, 97)_ = 44.78, *P* < 0.0001; PER2: Welch’s ANOVA, W (DFn, DFd) = 65.90 (2.000, 62.63), *P* < 0.0001; pCREB: Welch’s ANOVA, W (DFn, DFd) = 22.39 (2.000, 54.88), *P* < 0.0001; Figure 2).

**Figure 2.**
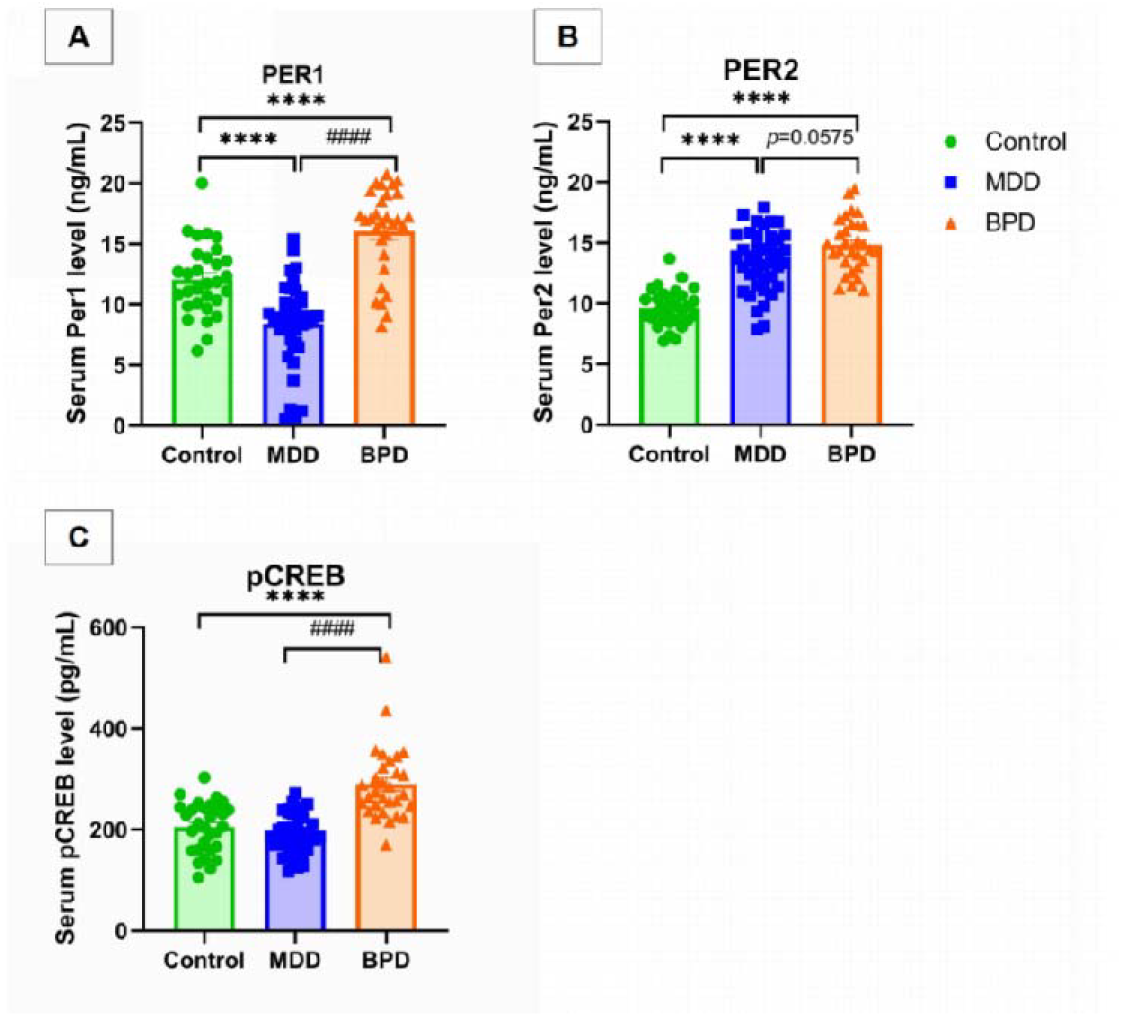
Serum levels of PER1, PER2 and pCREB in the control, MDD and BPD groups. **A**, Serum levels of PER1 detected by ELISA. Data were analyzed by One-way ANOVA with Tukey’s post-hoc test. *F* (2, 97) = 44.78, *P* < 0.0001; the tests on data normality and homogeneity of variance are shown in eFigure 2 in Supplement 1. **B**, Serum levels of PER2 detected by ELISA. Data were analyzed by Welch ANOVA tests with Dunnett’s T3 multiple comparisons test. W (DFn, DFd) = 65.90 (2.000, 62.63), *P* < 0.0001; the tests on data normality and homogeneity of variance are shown in eFigure 3 in Supplement 1. **C**, Serum levels of pCREB detected by ELISA. Data were analyzed by Welch ANOVA tests with Dunnett’s T3 multiple comparisons test. W(DFn, DFd) = 22.39 (2.000, 54.88), *P* < 0.0001. The tests on data normality and homogeneity of variance are shown in eFigure 4 in Supplement 1. ^****^*P* < 0.0001, compared with the control group; ^####^*P* < 0.0001, compared with the MDD group. Data were presented as mean ± SD. MDD, major depressive disorder; BPD, bipolar depression. Detailed data can be seen in eTable 2 in Supplement 1.

For further analysis by multiple comparisons, we found that compared with the HC group, serum PER1 levels were significantly reduced in the MDD group (*P* < 0.0001, Figure 2A), while serum PER2 levels were significantly increased in this group (*P* < 0.0001, Figure 2B). However, there were no significant differences in the pCREB levels in the MDD group vs the HC (Figure 2C). In the BPD group, serum levels of PER1, PER2 and pCREB were all significantly elevated (*P* < 0.0001; Figure 2), compared with the HCs. Both serum PER1 and pCREB levels were significantly higher in the BPD group than those in the MDD group (*P* < 0.0001; *P* < 0.0001, Figure 2A, C). However, there were only marginally significant difference in the serum PER2 levels between the two groups (*P* = 0.0575, Figure 2B). Therefore, serum PER1 and pCREB levels could be potentially used in differentiating MDD from BPD. The detailed data can be seen in eTable 2 and eFigure 2-4 in Supplement 1.

### Correlations between serum pCREB/PER1/PER2 levels and age/HAMD-17 score/MDQ score

To further explore whether serum levels of pCREB, PER1 and PER2 are correlated with the severity of MDD and BPD, we performed simple linear regression analyses. As shown in Figure 3 and eTable 3 in Supplement 1, we found that in the MDD group, serum PER1 levels were significantly positively related to MDQ scores (*r* = 0.364, *P* = 0.021, Figure 3C); however, the serum pCREB levels were significantly negatively related to MDQ scores (*r* = −0.338, *P* = 0.033, Figure 3F), thus indicating that serum PER1 and pCREB may be potential biomarkers for MDD.

**Figure 3.**
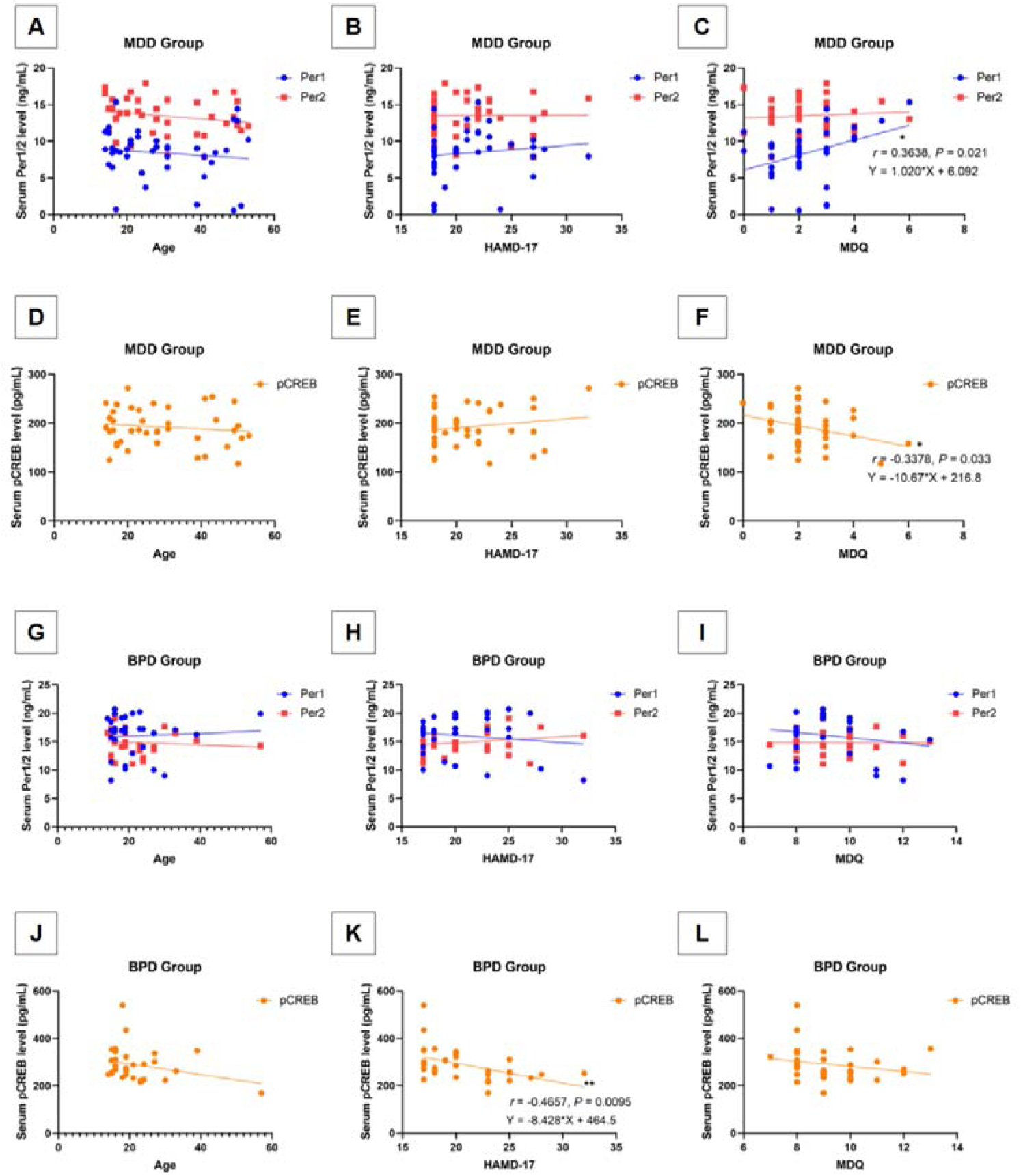
Correlation between serum PER1, PER2 and pCREB levels and Age/ HAMD/MDQ scores in the MDD and BPD groups. In the MDD group: **A**, Relationship between serum PER1/2 levels and Age. Age & PER1: Pearson *r* = −0.1187, *P* = 0.4656; Age & PER2: Pearson *r* = 0.1984, *P* = 0.2197; **B**, Relationship between serum PER1/2 levels and HAMD-17. HAMD-17 & PER1: Pearson *r* = 0.1241, *P* = 0.4456; HAMD-17 & PER2: Pearson *r* = 0.005742, *P* = 0.9719; **C**, Relationship between serum PER1/2 levels and MDQ. MDQ & PER1: Pearson *r* = 0.3638, \**P* = 0.0210; MDQ & PER2: Pearson *r* = 0.06683, *P* = 0.6820; **D**, Relationship between pCREB levels and Age: Pearson *r* = −0.1282, *P* = 0.4305; **E**, Relationship between pCREB levels and HAMD-17: Pearson *r* = 0.1786, *P* = 0.2703; **F**, Relationship between pCREB levels and MDQ: Pearson *r* = −0.3378, \**P* = 0.033. In the BPD group: **G**, Relationship between serum PER1/2 levels and Age. Age & PER1: Pearson *r* = 0.05647, *P* = 0.7669; Age & PER2: Pearson *r* = −0.08584, *P* = 0.6520; **H**, Relationship between serum PER1/2 levels and HAMD-17. HAMD-17 & PER1: Pearson *r* = −0.1679, *P* = 0.3752; HAMD-17 & PER2: Pearson *r* = 0.1808, *P* = 0.3389; **I**, Relationship between serum PER1/2 levels and MDQ. MDQ & PER1: Pearson *r* = −0.1929, *P* = 0.3071; MDQ & PER2: Pearson *r* = −0.006065, *P* = 0.9746; **J**, Relationship between pCREB levels and Age: Pearson *r* = −0.2802, *P* = 0.1336; **K**, Relationship between pCREB levels and HAMD-17: Pearson *r* = −0.4657, \*\**P* = 0.0095; **L**, Relationship between pCREB levels and MDQ: Pearson *r* = −0.2140, *P* = 0.2561. Data were analyzed by simple Pearson correlation analysis, \**P* < 0.05, \*\**P* < 0.01. MDD, major depressive disorder; BPD, bipolar depression; HAMD, Hamilton Depression Scale; MDQ, mood disorder questionnaire.

In the BPD group, serum pCREB levels were significantly negatively related to HAMD-17 scores (*r* = −0.4657, *P* = 0.0095, Figure 3K), thus demonstrating that serum pCREB is related to depressive symptoms of BD and may be a biomarker for BPD. However, we did not observe any significant relationship between age and serum pCREB, PER1 and PER2 levels in all groups (*P* > 0.05, Figure 3A, D, G and J; eFigure 1A and B; eTable 3 in Supplement 1).

In addition, there was no significant relationship between serum levels of PER1/PER2/pCREB and HAMD-17 scores in the MDD group (Figure 3B, E), and there was also no significant relationship between serum levels of PER1/PER2/pCREB and MDQ scores in the BPD group (Figure 3I, L; eTable 3 in Supplement 1), indicating that serum PER1/PER2/pCREB levels were not related to the severity of MDD.

### ROC analyses of serum pCREB, PER1 and PER2 in discriminating MDD from BPD

Next, we evaluated the role of serum PER1, PER2 and pCREB separately and jointly in differentiating MDD from BPD. We analyzed the sensitivity, specificity, Youden index and ROC curves of these potential biomarkers in the diagnosis and differential diagnosis of MDD and BPD. eTables 4-6 in Supplement 1 show the sensitivity, specificity, Youden index and the area under the ROC curve (AUC) for serum PER1, PER2 and pCREB in the diagnosis of MDD (eTable 4) and BPD (eTable 5), as well as the differential diagnosis between them (eTable 6). Moreover, Figure 4 shows the ROC curves for serum PER1, PER2 and pCREB in the diagnosis and differential diagnosis of MDD and BPD.

**Figure 4.**
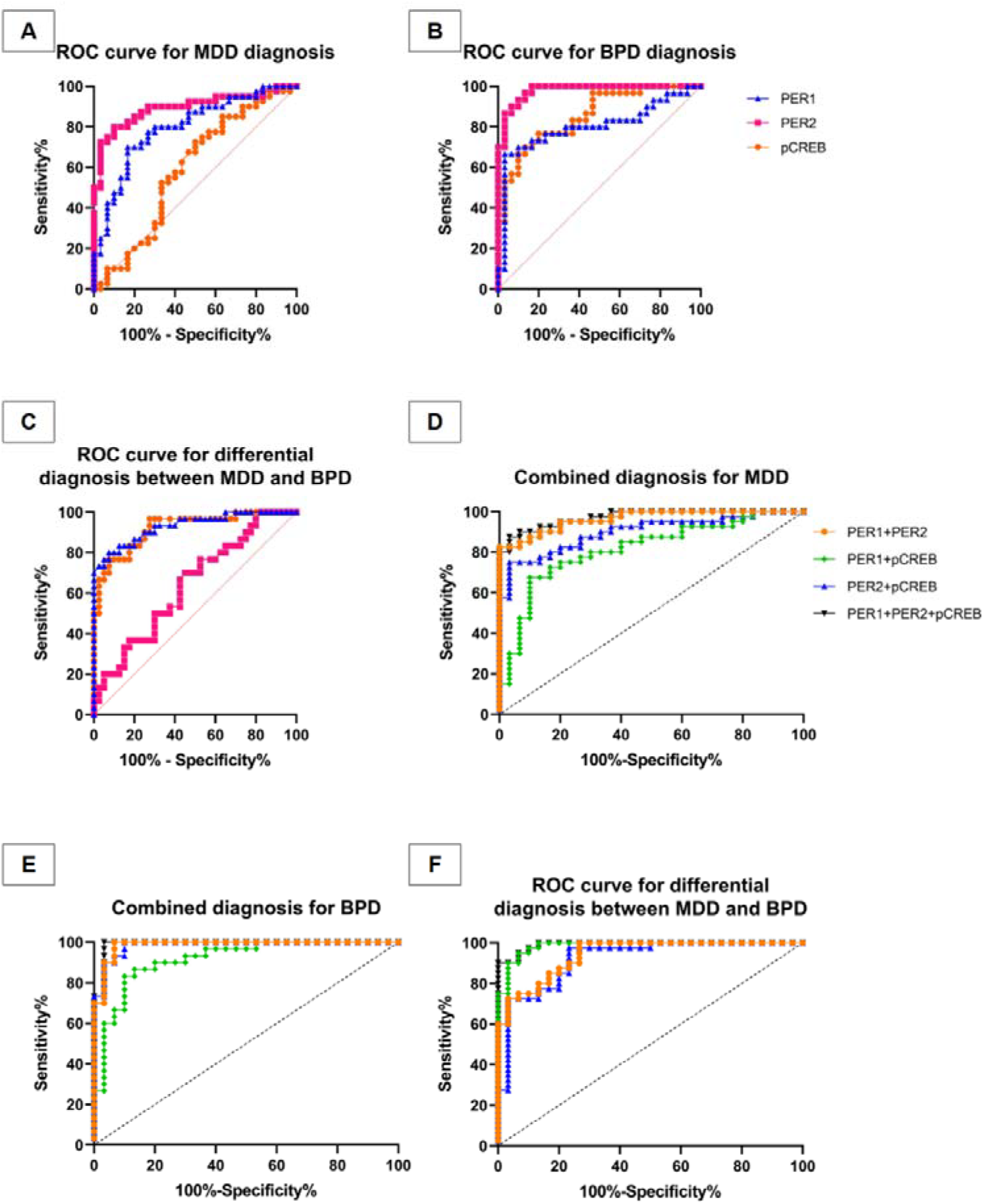
ROC curves for the respective or combined roles of serum PER1, PER2 and pCREB in the diagnosis and differential diagnosis of MDD and BPD. **A**, ROC curve for PER1, PER2 and pCREB in the diagnosis of MDD. **B**, ROC curve for PER1, PER2 and pCREB in the diagnosis of BPD. **C**, ROC curve for PER1, PER2 and pCREB in the differential diagnosis between BPD and MDD. **D**, ROC curve for combined biomarkers of PER1 & PER2, PER1 & pCREB, PER2 & pCREB, as well as PER1 & PER2 & pCREB in the diagnosis of MDD. **E**, ROC curve for combined biomarkers of PER1 & PER2, PER1 & pCREB, PER2 & pCREB, as well as PER1 & PER2 & pCREB in the diagnosis of BPD. **F**, ROC curve for combined biomarkers in the differential diagnosis between BPD and MDD. Detailed values can be seen in eTables 3-5. MDD, major depressive disorder; BPD, bipolar depression.

Regarding the diagnosis of MDD, we found that serum PER2 showed the highest diagnostic value among the three respective markers, as it yielded the highest sensitivity, specificity, Youden index and AUC (MDD vs HC, AUC = 0.8942; eTable 4 and Figure 4A). Multiple regression analysis revealed that the combined use of all the three biomarkers yielded an even higher sensitivity, Youden index, and AUC values (MDD vs HC, AUC = 0.9708; eTable 4 and Figure 4D), showing that serum PER2 may play important role in helping diagnose MDD.

Regarding the diagnosis of BPD, we found that serum PER2 also exhibited the highest diagnostic value among all the three biomarkers in the ROC analysis, as it yielded the highest sensitivity, Youden index and AUC values (BPD vs HC, AUC = 0.9789; eTable 5 and Figure 4B). Moreover, multiple regression analysis also showed the combined three biomarkers had even higher values in the diagnosis (BPD vs HC, AUC = 0.9911; eTable 5 and Figure 4E). However, this is only for differentiating BPD from HC, for the entire BD disease, the manic episode should also be taken into consideration, therefore, it’s still unclear whether PER2 is potential biomarker for BD.

Regarding differentiating MDD from BPD, we found that serum PER1 showed the highest diagnostic value among all the three biomarkers, as it yielded the highest specificity, Youden index and AUC values (MDD vs BPD, AUC = 0.9313; eTable 6 and Figure 4C). Combined use of serum PER1 and pCREB vs the combined three biomarkers yielded similarly high Youden index and AUC values (MDD vs BPD, AUC = 0.9858 and AUC = 0.9906 respectively; eTable 6 and Figure 4F). Therefore, combined use of serum PER1 and pCREB achieved great efficacy in differentiating MDD from BPD.

## Discussion

Recently, there have been several studies revealing biomarkers for discriminating MDD from BPD, including gut microbial signatures ^38^, metabolomic biomarker signatures ^39^, cortisol levels ^40^, proteomic profiling ^41^, as well as inflammatory biomarkers ^42^. To our knowledge, this is the first study into the role of circadian biomarker in the differential diagnosis between MDD and BPD, using cohorts and ELISA, we demonstrate that serum PER1 can be used in differentiating MDD and BPD, as it is decreased in the MDD group and increased in the BPD group; moreover, combined use of serum PER1/pCREB achieves similar high diagnostic efficacy in differentiating MDD from BPD as that of serum PER1/PER2/pCREB, with the sensitivity (0.933, 1), specificity (0.950, 0.897), Youden index (0.883, 0.897) as well as AUC (0.986, 0.991).

Previous studies have demonstrated that the sensitivity and specificity of the HAMD-17 for the diagnosis of MDD were 0.76 and 0.91, respectively ^43^; in addition, the Patient Health Questionnaire (PHQ-9) showed a sensitivity of 0.80 and a specificity of 0.92 for the diagnosis of MDD ^44^. In this study, we showed that the sensitivity and specificity of serum PER2 for the diagnosis of MDD were 0.8 and 0.9 respectively, similar as those of HAMD-17 and PHQ-9. While the combined three biomarkers showed a higher sensitivity and specificity with 0.875 and 0.967 separately, thus, showing higher diagnostic value for MDD than that of the above-mentioned scales.

In the differential diagnosis between MDD and BPD, previous study showed that the MDQ cutoff value is 7, with a sensitivity of 0.73 and a specificity of 0.90 ^37^. The cutoff value for the Hypomania Check List (HCL-32) has been found to be 13, with a sensitivity of 0.77 and a specificity of 0.62 ^45^. In addition, when the cutoff value of the YMRS was set at 29, the sensitivity was 0.83, and the specificity was 0.91; however, a score of 20 to 29 indicates a severe manic state and therefore has little significance for differentiating patients with a depressive state ^46^. Compared with the above scales, the combined serum PER1 and pCREB showed a higher sensitivity of 0.933 and a specificity of 0.950, a Youden index of 0.883 and AUC of 0.986, indicating great efficacy in differentiating MDD from BPD. Besides, in the MDD group, serum PER1 levels were significantly decreased compared with the HC group, consistent with previous animal study performed in the lateral habenula ^20^, on contrary, serum PER2 levels were significantly increased in the MDD group, compared with the HC group, which has not been demonstrated before, and we firstly revealed here that PER1 and PER2 may play opposite roles in MDD. However, serum pCREB levels was not significantly altered in this group versus the HC group, inconsistent with previous studies ^47, 48^, probably due to the limited sample size. We also found serum PER1 was positively related with MDQ scores, indicating that PER1 might be a risk factor to develop into BD, while serum pCREB was significantly negatively related to MDQ scores in patients with MDD, implicating that pCREB might be a maintaining factor for UPD, in agreement with previous studies ^49^.

In the BPD group, serum PER1, PER2 and pCREB levels were all significantly higher than the HC group, in agreement with previous studies ^32^. Among these, serum pCREB levels were significantly negatively related with HAMD-17 scores, thus may be related with the severity of BPD, consistent with previous studies ^33, 50^.

In differentiating MDD from BPD, we found pCREB and PER1 showed higher efficacy than PER2 respectively, probably because they are associated with the scores of clinical scales, however, PER2 showed low discriminating efficacy (*P* = 0.0575, Figure 2; sensitivity of 0.7, specificity of 0.55, Youden index of 0.25 and AUC of 0.64, eTable 6 in Supplement 1) and was not significantly related with any scale scores.

In addition, the average age of the MDD group was 7.62 years higher than that of the BPD group. There were no significant correlations between age and serum PER1, PER2, or pCREB levels in any of the three groups, thus indicating that age had no significant effect on the results. Previous studies have shown that the age of BD onset is younger than the age of MDD onset, and the risk of unipolar depression transitioning into bipolar depression decreases with age ^2^. Several studies have revealed that age has a minimal impact on serum levels of PER1/PER2 ^51, 52^, in line with our results.

Due to the limitations of case-control study, we examined risk factors for MDD and BPD to discriminate them, in the future, we may conduct cohort studies before and after treatments to explore the changes of these biomarkers in the progression of diseases.

## Conclusions

This differential diagnostic study demonstrated serum PER1 as a biomarker for discriminating MDD from BPD, and that combined use of serum pCREB and PER1 as efficient biomarkers, similar as combined use of pCREB, PER1 and PER2 in the differential diagnosis of MDD and BPD. In the future, it’s promising to explore whether they can be used to evaluate clinical treatment effects.

## Data Availability

The original data of this project is available upon reasonable request from the corresponding author, Dr. Wei Chen.

## Author contributions

Wang XL conceived this project, wrote the manuscript and provided a small part of the fund, Huang L performed the experiment and the statistical analysis, and they contributed equally to the paper; Yao JS, Qin YH and Ren KM provided the clinical cases and performed the statistical analysis, Shen YD and Chen W conceived this project, provided the clinical cases, applied for the funds and supervised this process, and they contribute equally to the manuscript.

## Data Sharing Statement

See Supplement 2.

## Supplementary Information

The supplementary materials can be available in the Supplement 1 and Supplement 2.

