## Supplemental Table 1-6 and Supplemental Figure 1-4 for "Circadian biomarker signatures for differentiating unipolar from bipolar depression": Supplement 1.docx

**Supplemental Online Content**

**eTable 1.** Comparison of basic demographic data

**eTable 2.** Comparisons of serum PER1, PER2 and pCREB levels (related to Figure 2)

**eTable 3.** Parameters for the correlation analysis between serum PER1, PER2 and pCREB and Age/ HAMD-17/MDQ (related to Figure 3)

**eTable 4.** Parameters for the serum PER1, PER2 and pCREB in the diagnosis of MDD (related to Figure 4)

**eTable 5.** Parameters for the serum PER1, PER2 and pCREB in the diagnosis of BPD (related to Figure 4)

**eTable 6.** Parameters for the serum PER1, PER2 and pCREB in the differential diagnosis between MDD and BPD (related to Figure 4)

eFigure 1. Correlation between serum pCREB, PER1 and PER2 levels and Age in the control group.

eFigure 2. Normality and homogeneity of variance tests for the data of PER1 in Figure 2

eFigure 3. Normality and homogeneity of variance tests for the data of PER2 in Figure 2

eFigure 4. Normality and homogeneity of variance tests for the data of pCREB in Figure 2

**eTable 1.** Comparison of basic demographic data

|  | **Health Control** | | | | **MDD** | | | | | **BPD** | | | | |  |
| --- | --- | --- | --- | --- | --- | --- | --- | --- | --- | --- | --- | --- | --- | --- | --- |
|  | **Mean** | **SD** | **Min** | **Max** | **Mean** | **SD** | **Min** | **Max** | ***P^a^*** | **Mean** | **SD** | **Min** | **Max** | ***P^b^*** | ***P^c^*** |
| *n* | 30 | | | | 40 | | | |  | 30 | | | |  |  |
| Sex | 21 female, 9 male | | | | 31 female, 9 male | | | |  | 19 female, 11 male | | | |  |  |
| Age | 30.43 | 8.52 | 18 | 55 | 29.25 | 12.91 | 14 | 53 | 0.955 | 21.63 | 8.95 | 14 | 57 | 0.001 | 0.014 |
| Education | 13.17 | 2.85 | 6 | 16 | 12.33 | 3.46 | 6 | 18 | 0.515 | 11.63 | 3.03 | 6 | 20 | 0.151 | 0.638 |
| HAMD-17 | 0.13 | 0.43 | 0 | 2 | 21.05 | 3.64 | 18 | 32 | 0.000 | 20.70 | 3.98 | 18 | 32 | 0.000 | 0.974 |
| MDQ | 0.63 | 0.72 | 0 | 2 | 2.28 | 1.22 | 0 | 6 | 0.000 | 9.30 | 1.42 | 7 | 13 | 0.000 | 0.000 |

Data are shown as mean ± SD. Age, scores of HMAD-17 and MDQ were analyzed by Welch ANOVA with Dunnett's T3 multiple comparisons test. Besides, education duration was analyzed by One-way ANOVA with Tukey’s multiple comparisons test. *F_age_*=6.620; *F_education_*=1.770; *F_HAMD-17_*=447.54; *F_MDQ_* =473.400. *P*^a^, MDD vs. Health control group; *P*^b^, BPD vs. Health control group; *P*^c^, BPD vs. MDD group.

**eTable 2.** Comparisons of serum PER1, PER2 and pCREB levels (related to Figure 2)

|  | **Control** | **MDD** | | **BPD** | | **MDD vs BPD** |  |  |
| --- | --- | --- | --- | --- | --- | --- | --- | --- |
|  | **Mean ± SD** | **Mean ± SD** | **Adjusted *P*^a^** | **Mean ± SD** | **Adjusted**  ***P*^b^** | **Adjusted *P*^c^** | ***F*** | **Test** |
| PER1  (ng/mL) | 12.05±2.96 | 8.41±2.96 | **** | 16.05±3.60 | **** | **** | *F* _(2, 97)_ = 44.78 | One-way ANOVA, with Tukey’s multiple comparisons test |
| PER2  (ng/mL) | 9.67±1.54 | 13.51±2.49 | **** | 14.86±2.22 | **** | 0.0575 | W _(DFn, DFd)_ = 65.90 (2.000, 62.63) | Welch ANOVA, Dunnett's T3 multiple comparisons test |
| pCREB  (pg/mL) | 205.20±49.57 | 192.60±38.52 | ns | 290.00±72.10 | **** | **** | W _(DFn, DFd)_ = 22.39 (2.000, 54.88) | Welch ANOVA, Dunnett's T3 multiple comparisons test |

Adjusted *P*^a^, comparison between the MDD and control group; Adjusted *P*^b^, comparison between the BPD and control group; Adjusted *P*^c^, comparison between the MDD and BPD group. *****P* < 0.0001.

**eTable 3.** Parameters for the correlation analysis between serum PER1, PER2 and pCREB and Age/ HAMD-17/MDQ (related to Figure 3 and eFigure 1)

| **Pearson *r*, *P* (two-tailed)** | | **Age** | **HAMD-17** | **MDQ** |
| --- | --- | --- | --- | --- |
| **MDD** | **PER1** | *r* = -0.1187, *P* = 0.4656 | *r* = 0.1241, *P* = 0.4456 | *r* = 0.3638, ******P* = 0.0210 |
|  | **PER2** | *r* = -0.1984, *P* = 0.2197 | *r* = 0.0057, *P* = 0.9719 | *r* = 0.0668, *P* = 0.6820 |
|  | **pCREB** | *r* = -0.1282, *P* = 0.4305 | *r* = 0.1786, *P* = 0.2703 | *r* = -0.3378, ******P* = 0.0330 |
| **BPD** | **PER1** | *r* = 0.0565, *P* = 0.7669 | *r* = -0.1467, *P* = 0.4393 | *r* = -0.1929, *P* = 0.3071 |
|  | **PER2** | *r* = -0.0858, *P* = 0.6520 | *r* = 0.1972, *P* = 0.2964 | *r* = -0.0061, *P* = 0.9746 |
|  | **pCREB** | *r* = -0.2802, *P* = 0.1336 | *r* = -0.4657, *******P* = 0.0095 | *r* = -0.2140, *P* = 0.2561 |

**eTable 4.** Parameters for the serum PER1, PER2 and pCREB in the diagnosis of MDD (related to Figure 4)

|  | **Sensitivity**  **(95%CI)** | **Specificity**  **(95%CI)** | **Youden index** | **AUC**  **(95%CI)** |
| --- | --- | --- | --- | --- |
| PER1 | 0.700（0.546-0.819） | 0.833（0.664-0.926） | 0.533 | 0.7983 (0.6933-0.9034) |
| PER2 | 0.800（0.652-0.895） | 0.900（0.743-0.965） | 0.700 | 0.8942 (0.8175-0.9709) |
| pCREB | 0.725（0.518-0.839） | 0.500（0.332-0.669） | 0.225 | 0.5892 (0.4483-0.7300) |
| PER1+PER2 | 0.825（0.681-0.913） | 1.000（0.887-1.000） | 0.825 | 0.9633 (0.9271-0.9996) |
| PER1+pCREB | 0.675（0.520-0.800） | 0.900（0.774-0.965） | 0.575 | 0.8150 (0.7131-0.9169) |
| PER2+pCREB | 0.750（0.598-0.858） | 0.967（0.833-0.998） | 0.717 | 0.9000 (0.8287-0.9713) |
| PER1+PER2+pCREB | 0.875（0.739-0.945） | 0.967（0.833-0.998） | 0.841 | 0.9708 (0.9396-1.000) |

The data were analyzed by ROC analysis for one biomarker, binary regression analysis for two combined biomarkers and multiple regression analysis for three biomarkers.

**eTable 5.** Parameters for the serum PER1, PER2 and pCREB in the diagnosis of BPD (related to Figure 4)

|  | **Sensitivity (95%CI)** | **Specificity**  **(95%CI)** | **Youden index** | **AUC (95%CI)** |
| --- | --- | --- | --- | --- |
| PER1 | 0.667（0.488-0.808） | 0.967（0.833-0.998） | 0.634 | 0.8033 (0.6837-0.9229) |
| PER2 | 0.967（0.833-0.998） | 0.867（0.703-0.947） | 0.834 | 0.9789 (0.9514-1.000) |
| pCREB | 0.767（0.591-0.882） | 0.800（0.627-0.945） | 0.567 | 0.8550 (0.7623-0.9477) |
| PER1+PER2 | 1.000（0.887-1.000） | 0.933（0.787-0.988） | 0.933 | 0.9867 (0.9641-1.000) |
| PER1+pCREB | 0.867（0.703-0.947） | 0.867（0.703-0.947） | 0.734 | 0.9167 (0.8458-0.9875) |
| PER2+pCREB | 1.000（0.887-1.000） | 0.900（0.744-0.965） | 0.900 | 0.9856 (0.9635-1.000) |
| PER1+PER2+pCREB | 1.000（0.887-1.000） | 0.967（0.833-0.998） | 0.967 | 0.9911 (0.9725-1.000) |

The data were analyzed by ROC analysis for one biomarker, binary regression analysis for two combined biomarkers and multiple regression analysis for three biomarkers.

**eTable 6.** Parameters for the serum PER1, PER2 and pCREB in the differential diagnosis between MDD and BPD (related to Figure 4)

|  | **Sensitivity (95%CI)** | **Specificity**  **(95%CI)** | **Youden index** | **AUC (95%CI)** |
| --- | --- | --- | --- | --- |
| PER1 | 0.800（0.627-0.905） | 0.925（0.801-0.974） | 0.725 | 0.9313 (0.8715-0.9910) |
| PER2 | 0.700（0.521-0.833） | 0.550（0.398-0.693） | 0.250 | 0.6404 (0.5109-0.7699) |
| pCREB | 0.967（0.833-0.998） | 0.725（0.572-0.839） | 0.692 | 0.9196 (0.8554-0.9838) |
| PER1+PER2 | 0.733（0.556-0.858） | 1.000（0.912-1.000） | 0.733 | 0.9417 (0.8925-0.9908) |
| PER1+pCREB | 0.933（0.787-0.988） | 0.950（0.744-0.965） | 0.883 | 0.9858 (0.9654-1.000) |
| PER2+pCREB | 0.767（0.591-0.882） | 0.975（0.871-0.999） | 0.742 | 0.9217 (0.8570 -0.9863) |
| PER1+PER2+pCREB | 1.000（0.887-1.000） | 0.897（0.764-0.960） | 0.897 | 0.9906 (0.9764-1.000) |

The data were analyzed by ROC analysis for one biomarker, binary regression analysis for two combined biomarkers and multiple regression analysis for three biomarkers.

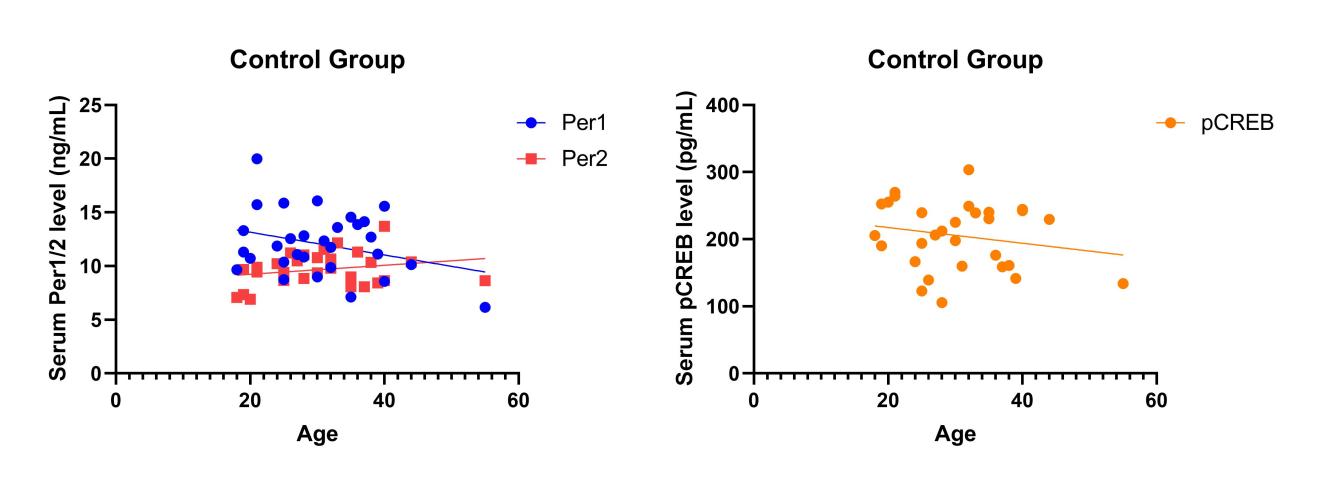

**B**

**A**

**eFigure 1.** Correlation between serum pCREB, PER1 and PER2 levels and Age in the control group

**A**, Relationship between serum PER1/2 levels and Age. Age vs. PER1: Pearson *r* = -0.3052, *P* = 0.1010; Age vs. PER2: Pearson *r* =0.2321, *P* = 0.2171; **B**, Relationship between pCREB levels and Age: Pearson *r* = -0.2009, *P* = 0.2871. Pearson correlation analysis, *P* > 0.05.

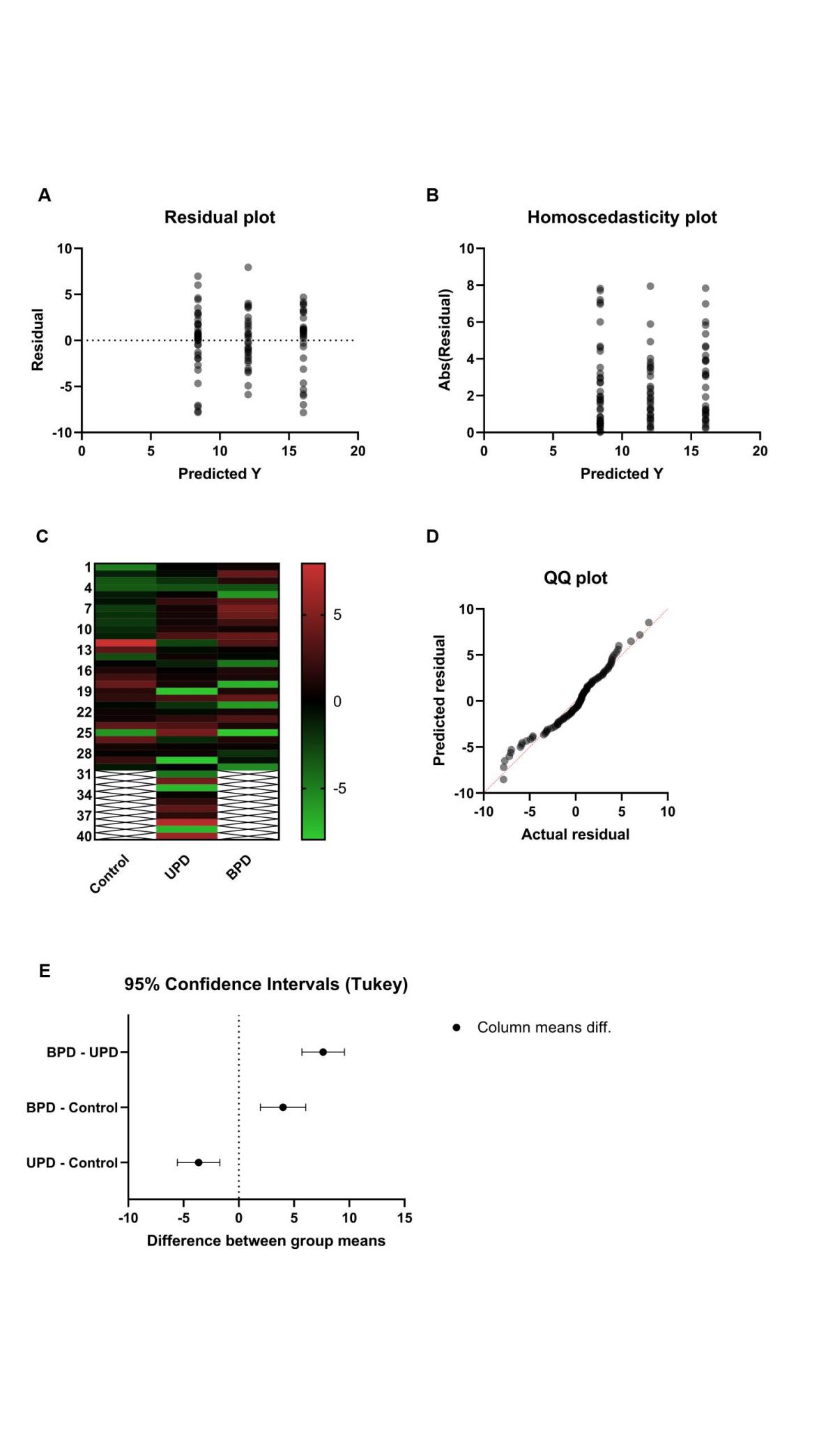

**eFigure 2.** Normality and homogeneity of variance tests for the data of PER1 in Figure 2

**A**, Residual plot, **B**, Homoscedasticity plot, **C**, Heat map plot, **D**, QQ plot, **E**, Multiple comparisons’ CI plot.

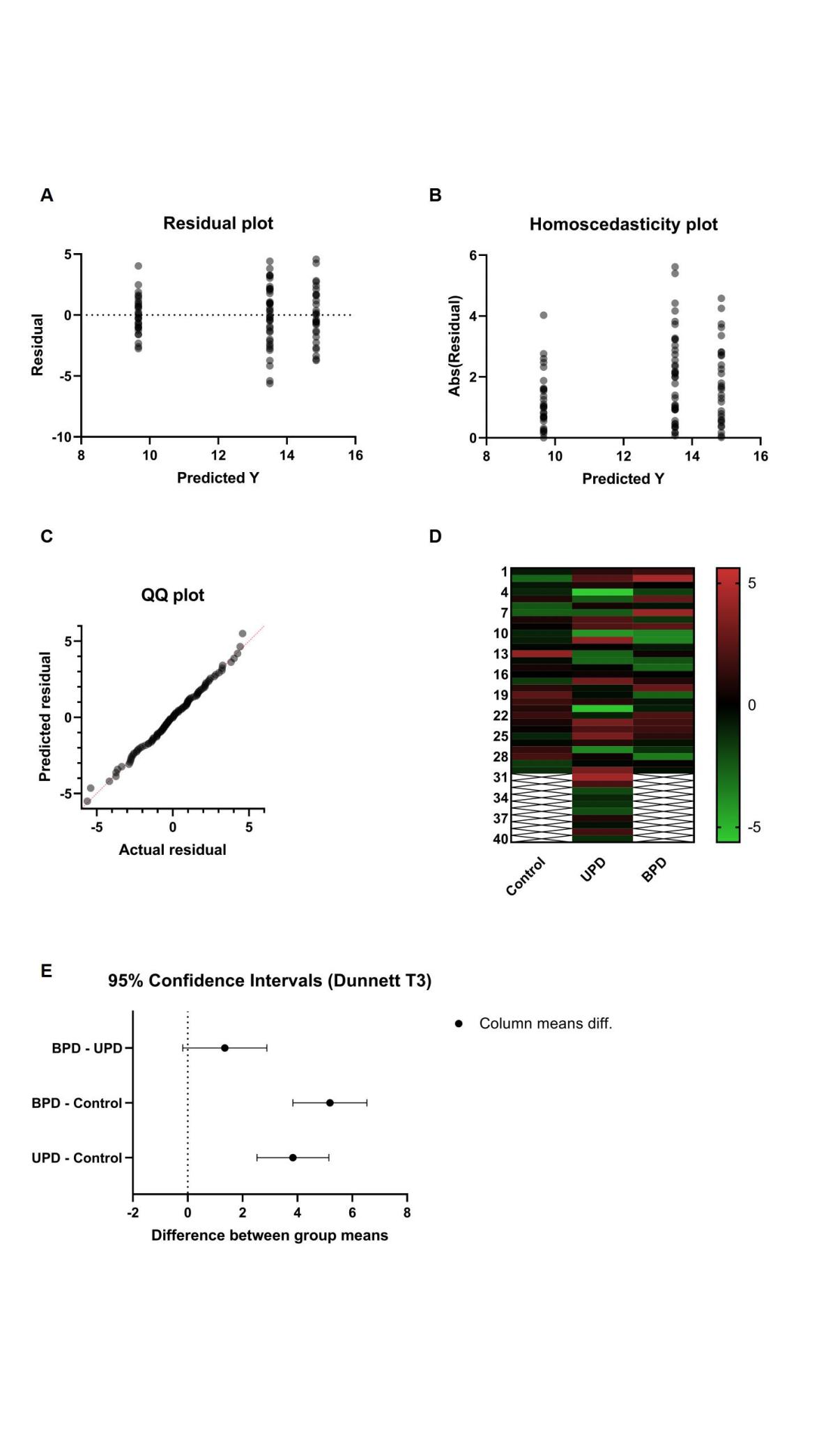

**eFigure 3.** Normality and homogeneity of variance tests for the data of PER2 in Figure 2

**A**, Residual plot, **B**, Homoscedasticity plot, **C**, QQ plot, **D**, Heat map plot, **E**, Multiple comparisons’ CI plot.

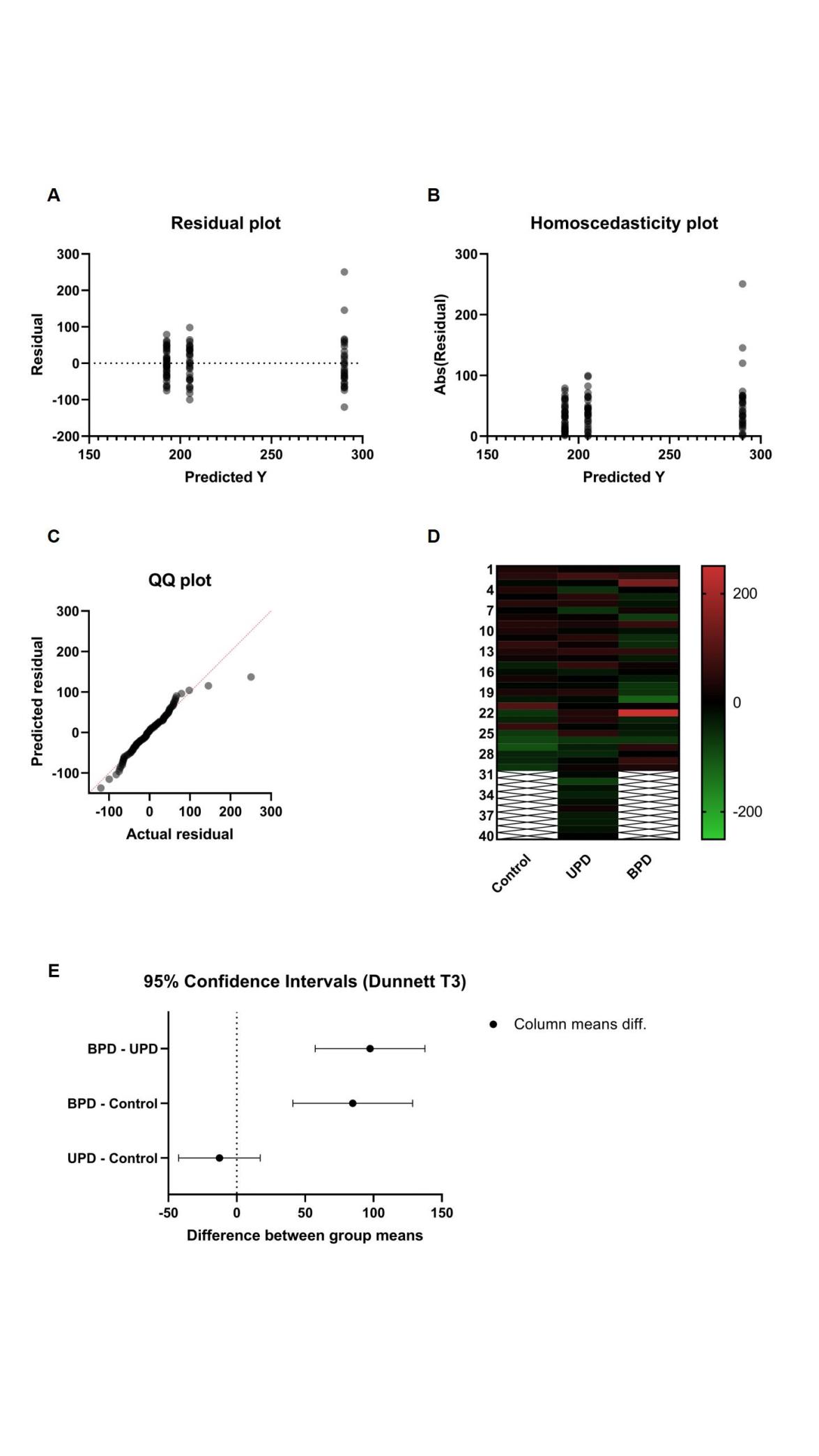

**eFigure 4.** Normality and homogeneity of variance tests for the data of pCREB in Figure 2

**A**, Residual plot, **B**, Homoscedasticity plot, **C**, QQ plot, **D**, Heat map plot, **E**, Multiple comparisons’ CI plot.
